# Daily variation in blood glucose levels during continuous enteral nutrition in patients on the Intensive Care Unit: a retrospective observational study

**DOI:** 10.1101/2023.10.04.23296529

**Authors:** Floor W. Hiemstra, Dirk Jan Stenvers, Andries Kalsbeek, Evert de Jonge, David J. van Westerloo, Laura Kervezee

## Abstract

**Background:** The circadian timing system coordinates daily cycles in physiological functions, including glucose metabolism and insulin sensitivity. Here, the aim was to characterize the 24-hour variation in glucose levels in critically ill patients during continuous enteral nutrition after controlling for potential sources of bias.

**Methods:** Time-stamped clinical data from adult patients who stayed in the Intensive Care Unit (ICU) for at least 4 days and received enteral nutrition were extracted from the Medical Information Mart for Intensive Care (MIMIC)-IV database. Linear mixed-effects and XGBoost modelling were used to determine the effect of time of day on blood glucose values.

**Findings:** In total, 207,647 glucose measurements collected during enteral nutrition were available from 6,929 ICU patients. Using linear mixed-effects modelling, time of day had a significant effect on blood glucose levels (p<0·001), with a peak of 9·6 [9·5 – 9·6; estimated marginal means, 95% CI] mmol/L at 10:00 in the morning and a trough of 8·6 [8·5 – 8·6] mmol/L at 03:00 at night. A similar impact of time of day on glucose levels was found with the XGBoost regression model.

**Interpretation:** These results revealed marked 24-hour variation in glucose levels in ICU patients even during continuous enteral nutrition. This 24-hour pattern persists after adjustment for potential sources of bias, suggesting it may be the result of endogenous biological rhythmicity.

**Funding:** This work was supported by a VENI grant (2020-09150161910128) from the Netherlands Organization for Health Research and Development (ZonMw), an institutional project grant, and by the Dutch Research Council (NWO).

## Introduction

The circadian timing system coordinates daily cycles in physiological functions, including glucose metabolism.^1^ Local tissue clocks work in concert with the central circadian clock in the suprachiasmatic nucleus to optimally prepare the body for the predictable daily variations in feeding and fasting and rest and activity. As such, physiological processes affecting plasma glucose levels, such as endogenous glucose production, glucose-induced insulin secretion, and local insulin sensitivity all show marked 24-hour rhythms.^2^ In healthy humans, this results in a lower increase in glucose levels in response to food intake in the morning as compared to in the evening and night. In contrast, in people with diabetes mellitus, these 24-hour rhythms give rise to the so called dawn phenomenon; a sharp rise in blood glucose levels in the early morning as a result of reduced insulin sensitivity at the time of highest endogenous glucose production in the early morning.^3^

The relevance of these daily changes in glucose regulation in real-world settings is exemplified by various lines of evidence. For example, a large-scale study in which adults not known to have diabetes were randomized to have their blood glucose examined in either the morning or the afternoon.^4^ Despite a shorter fasting duration for afternoon blood draws (breakfast was allowed), fasting glucose levels were lower in the afternoon than in the morning. It was estimated that half of the diabetes cases would go undiagnosed in the afternoon group with the current diagnostic criteria, underscoring that time of blood sampling is an important consideration in the diagnosis of diabetes mellitus specifically, and in clinical practice in general.

In hospitalized patients, various outcomes related to glucose regulation have also been reported to vary by time of day. In general, several retrospective studies have found that low glucose levels occur more frequently at night compared to other times of day in adult inpatients, both in those with diabetes mellitus^5–7^ and without diabetes mellitus.^8^ In addition, various retrospective studies have shown that blood glucose levels vary by time of day in critically ill patients in the Intensive Care Unit (ICU), with higher levels during the day and lower levels at night.^9–11^ Indeed, the importance of clock time for the prediction of glucose values is highlighted by the finding that time of day emerged as the third most important feature to forecast glucose values in ICU patients in a machine learning algorithm based on more than 70 clinical features derived from electronic health records.^12^

The origin of this daily variation in glucose control in hospitalized patients is unknown. On the one hand, some authors argued that this effect can be ascribed to 24-hour variation in underlying physiological processes, such as circadian changes in insulin sensitivity.^9^ On the other hand, this time-of-day dependent effect has mostly been attributed to daily variations in health care processes rather than physiology, such as daily changes in feeding support or exogenous insulin administration.^10,11^ Moreover, selection bias can play a role, as glucose levels are examined more frequently in patients with poor glucose control, while sampling in more stable patients may be restricted to a routine check during clinical rounds in the morning.^13^ Indeed, controlling for uneven sampling by using time-weighted average glucose levels was found to diminish the 24-hour variation in glucose levels in ICU patients.^13^ Clearly, identifying and adjusting for sources of bias is an essential step in the analysis of circadian patterns in electronic health record data. This is underlined by the finding that the hour of the day a laboratory test is ordered in the hospital can be more predictive for survival than the actual value of the test result.^14^

Thus, it remains unexplored to what extent blood glucose levels show 24-hour variation in clinical settings after controlling for the various sources of bias introduced by daily rhythms in clinical practice. Therefore, the goal of this study was to characterize 24-hour variation in blood glucose levels in critically ill patients during periods of continuous enteral tube feeding and to evaluate the effect of potential sources of bias that may influence these results. To this end, we made use of the Medical Information Mart for Intensive Care-IV (MIMIC-IV) database, containing detailed time-stamped, annotated clinical data of > 50,000 adult ICU patients, providing a wealth of data to study daily patterns in physiology in a real-world critical care setting. As such, this study aims to shed light on the daily dynamics of blood glucose regulation in critically ill patients, facilitating a deeper understanding of their daily physiological patterns.

## Methods

### Data source

Data were obtained from the MIMIC-IV (version 2.0) database.^15,16^ This is a publicly available, single-center critical care database consisting of detailed, time-stamped clinical data from over 50,000 patients ≥ 18 years and admitted to the ICU of the Beth Israel Deaconess Medical Center (Boston, Massachusetts) between 2008 and 2019. The database includes patient demographics and characteristics, measurements, procedures, administered medication and treatments sourced from the electronic health record. Two authors (FWH and LK) obtained access to the database by completing the Collaborative Institutional Training Initiative program and were responsible for data extraction and analysis (certification numbers: 45894318 and 35567467, respectively). The study was reported in accordance with the Strengthening the Reporting of Observational studies in Epidemiology (STROBE) statement^17^ and the REporting of studies Conducted using Observational Routinely collected health Data (RECORD) statement.^18^ The code of data extraction and analysis is available on GitHub (https://github.com/fwhiemstra/mimic-glucose-24h-variation-in-icu).

### Data extraction and pre-processing

Data from the MIMIC-IV database were extracted using structured query language within PostgreSQL (version 16, PostgreSQL Global Development Group) and pre-processing was performed using R programming language (version 4.3.1). Patients and ICU stays were selected from the database using the following inclusion criteria: 1) total length of ICU stay (from admission to discharge) ≥ 4 days; 2) ICU stay was not a readmission; 3) glucose measurements were taken during the ICU stay; and 4) enteral nutrition was administered during the ICU stay. All glucose measurements that were taken while patients received enteral nutrition during their ICU stay were extracted. Both point-of-care capillary tests as well as laboratory (serum and whole arterial blood) measurements were included. It was previously shown in the MIMIC-III database that the systematic bias between these measurement methods is limited.^19^ In the case of simultaneous glucose measurements, whole blood measurements were preferred over serum measurements, that were in their turn preferred over point-of-care capillary measurements, following a similar approach as Robles Arévalo et al.^19^ Glucose measurements with a value > 55·6 mmol/L (1,000 mg/dL) were excluded from the dataset, as they exceed the limit of accuracy for the laboratory analyzer that was used in the Beth Israel Deaconess Medical Center.^19^ For capillary samples, measurements with a value > 27·8 mmol/L (500 mg/dL) were excluded, according to the limit of accuracy for the point-of-care glucometers (Precision Xceed, Abbott Laboratories, Chicago, Illinois, United States). The charting time was used to time-stamp the glucose measurements as this is considered in the MIMIC-IV database to be the closest proxy to the time the data was actually measured. However, in case the ‘store time’ (the time the measurement was recorded by the care giver) was recorded before the ‘chart time’ (the time the actual data entry occurred), the ‘store time’ was used as time-stamp for the glucose measurement, as described previously.^19^ Time of day was rounded down to the nearest integer hour on a scale from 0 to 23.

For each patient, the following variables were extracted: age, sex, diagnosis of diabetes, in-hospital mortality, type of admission (elective or emergency), race, Charlson Comorbidity Index, Sequential Organ Failure Assessment (SOFA) score and Oxford Acute Severity of Illness Score (OASIS) score.^20–22^. Sample-level variables that vary with the timing of the glucose sampling within an individual ICU stay, were collected and time-matched to each glucose measurement in the dataset (administration of carbohydrate via nutrition, insulin, dextrose and glucocorticoids, type of mechanical ventilation and Richmond Agitation-Sedation Scale (RASS) score). Insulin and dextrose administrations were quantified (units/hour and grams/hour, respectively). Glucocorticoid administration was used as binary variables (yes/no). Rate of carbohydrate administration (grams per hour) was determined for each enteral nutrition event by multiplying the hourly nutrition rate by the carbohydrate content of the respective nutrition product (for details, see Supplementary Table S1). Glucose measurements that were taken during enteral nutrition events with unknown carbohydrate content were removed from the dataset. Continuous variables (age and administration rates of carbohydrates, dextrose and insulin) were categorized for use in the linear mixed-effects model. Boundaries for categorization were based on data distributions and can be viewed in Supplementary Table S2.

Next, time corrections were applied to the start and end time of the insulin administrations before matching the insulin events to the glucose measurements, in order to take into account the variation in the start and duration of action between different types of insulin. Start times of insulin administrations were corrected by adding 0.5 hours and end times were extended, depending on their mode of actions, with 2, 4, 10 and 12 hours for rapid-, short-, intermediate and long-action insulin, respectively.^19^ Additionally, a time correction was used for the administration of glucocorticoids by adding 24 hour to the end time of administration, thereby accounting for the sustained effects of glucocorticoids on glucose metabolism. Similarly, end times of drug push or bolus administrations of dextrose were prolonged to 10 minutes after start time. In the subsequent sections of this article, the glucose measurements taken during administration of insulin, dextrose and glucocorticoids, include those that fall within the time correction windows. Finally, days since ICU admission and time to the next glucose measurement (using all glucose measurements from the MIMIC-IV dataset, including those not collected during enteral nutrition) were derived for each glucose measurement in the data. An overview of the extracted variables is provided in Supplementary Figure S1 and Supplementary Table S2.

### Statistical analysis

Statistical analysis and visualization was performed using R programming language (version 4.3.1). To visualize variation of glucose levels over time, the glucose levels were normalized per patient and plotted against time. Next, a linear mixed-effects model was used to assess the effect of time of day on glucose levels, adjusted for relevant variables (LME4 package, version 1.1.30).^23^ Glucose levels (in mmol/L) were used as dependent variable and patient IDs were added as random effect on the intercept. Four different linear mixed-effects models were fitted, with each subsequent model incorporating additional fixed effects to evaluate the effect of adding fixed effects to the model (Supplementary Table S2). In model 1, only an intercept was used. In model 2, patient-level variables were added (age (categorized), sex, diabetes mellitus diagnosis (yes/no)) as fixed effects, followed by subsequent addition of sample-level variables in model 3 (carbohydrate administration rate (categorized), dextrose administration rate (categorized), insulin administration rate (categorized) and glucocorticoid administration (binary)). Selection of these fixed effects was based on their known, direct effect on glucose levels (Supplementary Figure S1). Finally, in model 4, in addition to the patient-level and sample-level variables, time (in hourly bins) was included as fixed effect. The models were fitted using the entire glucose data set, employing the Restricted Maximum Likelihood approach to estimate the variance of random effects, and subsequently subjected to likelihood ratio tests for model comparison. A significance level of 0.05 was considered as statistically significant and was used to select the final model. Estimated marginal means were calculated with the Maximum Likelihood approach and used to visualize the glucose levels over the 24-hour period. Residual plots were created for the final linear mixed-effects model to assess whether the model assumptions of linearity and constant variance were met.

As an alternative approach, the same dataset was analyzed with the decision tree-based machine learning method Extreme Gradient Boosting (XGBoost) to assess the time of day effect using Shapley Additive exPlanations (SHAP) values.^24,25^ In contrast to the linear mixed-effects models, the variables age and administration rate of carbohydrates, dextrose and insulin were not categorized and used as continuous variables (Supplementary Table S2). A detailed description of this method is available in the Supplementary Methods S1.

Next, sensitivity analyses were performed on various subgroups, based on variables that are considered to have an indirect effect on glucose levels or that may potentially bias the time-of-day dependent effect: type of mechanical ventilation, RASS scores (both as proxy for no oral intake), in-hospital mortality, days in ICU (both as proxy for severity of illness), sample type and sampling frequency (to rule out sampling bias) (Supplementary Figure S1). Furthermore, four subgroups were made to distinguish between patients without insulin requirement and with various levels of insulin requirements – who are thus assumed to have impaired glucose tolerance. Patients without insulin requirement were defined as those who did not receive insulin at any point during their ICU stay, and patients who required insulin therapy were divided in three groups: those who received an average daily administration of more than 0<x≤35, 35<x≤70, or > 70 units of insulin. For each subgroup, a linear mixed-effects model (using the same variables and settings as in model 4) was fitted. Estimated marginal means were used to visualize the 24-hour patterns for each of the subgroups.

## Results

### Baseline characteristics

A total of 6,929 ICU stays (52,333 ICU days) were included in the dataset, with a total of 207,647 valid glucose measurements that were taking during enteral nutrition (Figure 1). Baseline characteristics of the included patients and characteristics of the included glucose measurements are summarized in Table 1 and Supplementary Table S3, respectively. A representative example of the timing of enteral nutrition, glucose measurements, and the administration of dextrose, glucocorticoids, and insulin during a day in the ICU is shown in Figure 2A, and a typical pattern of enteral nutritional support across the ICU stay is shown in Figure 2B. Overall, the included glucose measurements were taken throughout the 24-hour period with an increase of the number of measurements at some time points, corresponding to the timing of routine lab rounds (Figure 2C). Time-of-day distributions of dosing rates of enteral nutrition and administrations of insulin, dextrose and glucocorticoids are shown in Figure 2D and Supplementary Figure S2, respectively.

**Figure 1:**
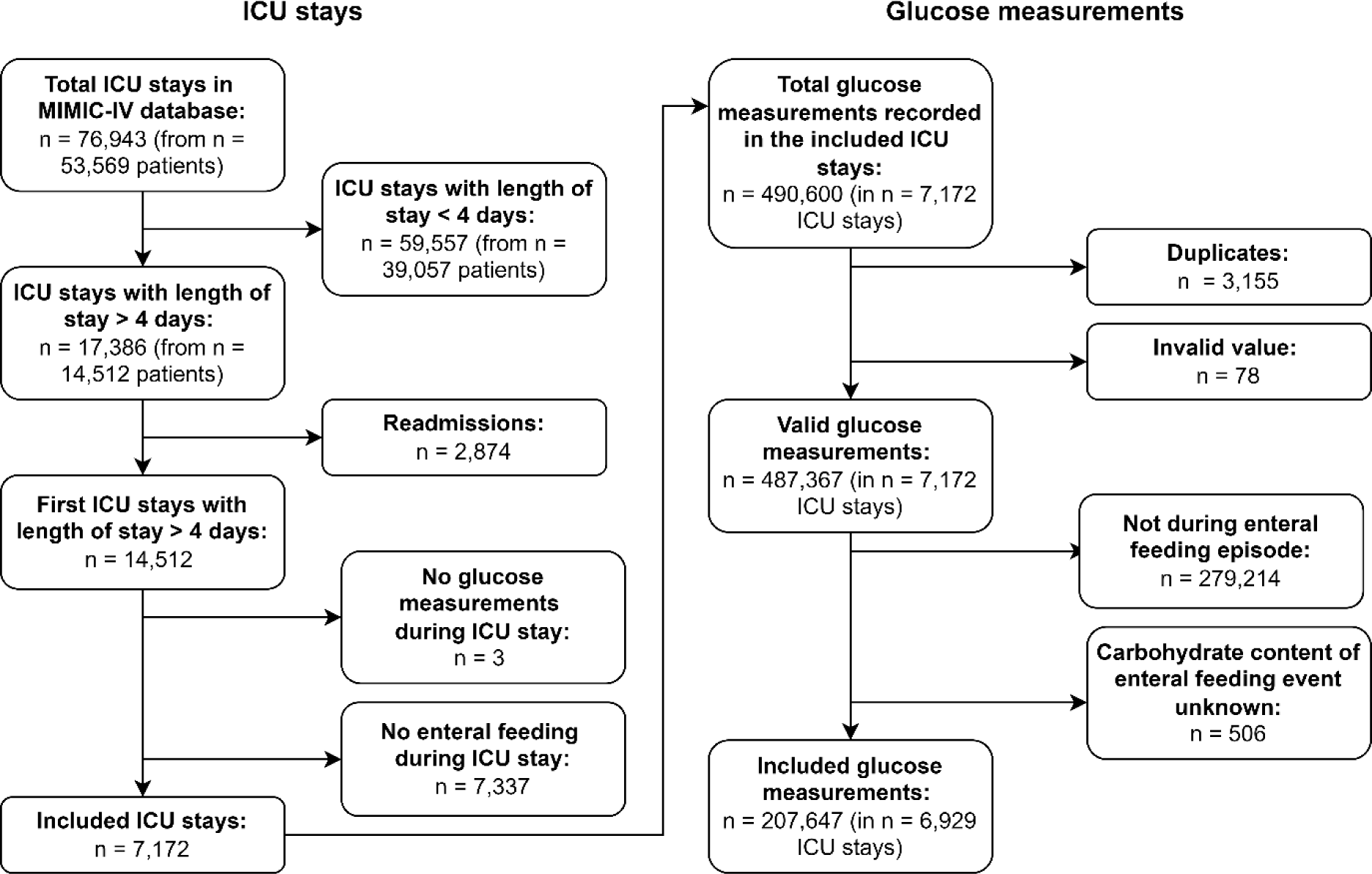
Inclusion flowchart of ICU stays and glucose measurements.

**Figure 2:**
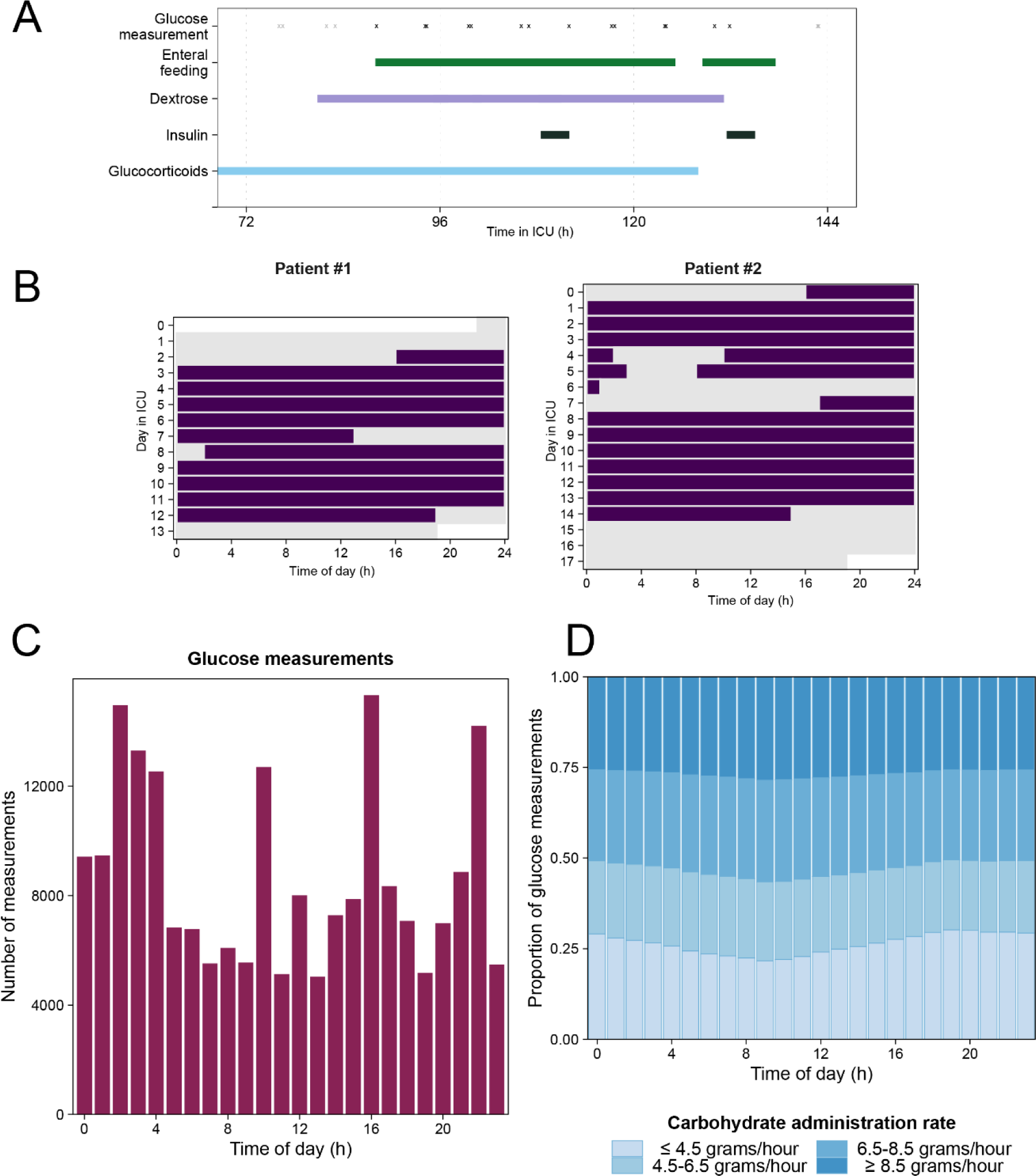
Distributions of glucose measurements and enteral nutrition rates over the 24-hour period and across the ICU stay. (**A**) Example of the timing of enteral nutrition, glucose measurements and administrations of dextrose, glucocorticoids and insulin in one representative patient. Black symbols represent glucose measurements taken while the patient received enteral nutritional support (included in the dataset), grey symbols represent glucose measurements taken while the patient did not receive enteral nutritional support (not included in the dataset). (**B**) Representative examples of enteral nutritional support across the ICU stay, taken from two patients included in the dataset. (**C**) Distribution of included glucose measurements, and (**D**) dosing rate of enteral nutrition (at the time of glucose measurements) by time of day.

**Table 1:** Baseline characteristics of included patients.

| **Patient characteristics** | **Patients (n=6,929)** |
| --- | --- |
| **Sex, n (%)** |  |
| **Male** | 3,948 (57%) |
| **Female** | 2,981 (43%) |
| **Age (years), median [IQR]** | 66 [54-77] |
| **Race, n (%)** |  |
| **White** | 4,245 (61%) |
| **Black** | 664 (9.6%) |
| **Hispanic** | 250 (3.6%) |
| **Asian** | 193 (2.8%) |
| **Other** | 270 (3.9%) |
| **Unknown** | 1,307 (19%) |
| **Admission type, n (%)** |  |
| **Elective** | 491 (7.1%) |
| **Emergency** | 6,438 (93%) |
| **SOFA score at admission, median [IQR]** | 8 [5-11] |
| **OASIS score, median [IQR]** | 40 [35-46] |
| **Length of stay (days), median [IQR]** | 10 [7-16] |
| **Diabetes diagnosis, n (%)** | 2,126 (31%) |
| **In-hospital mortality, n (%)** | 1,967 (28%) |
| **Mechanical ventilation during ICU stay, n (%)** |  |
| **Invasive ventilation** | 6,219 (90%) |
| **Non-invasive ventilation only** | 603 (8.7%) |
| **None / No ventilation data available** | 107 (1.5%) |

SOFA: Sequential Organ Failure Assessment<sup>22</sup>; OASIS: Oxford Acute Severity of Illness Score<sup>21</sup>; IQR: inter-quartile range

### Analysis of 24-hour variation in glucose levels during enteral nutrition

Without adjustment for potential confounders, normalized glucose levels showed substantial variation over time, with a peak at 10:00h and trough at 03:00h (Supplementary Figure S3). The linear mixed-effects model fits improved significantly each time new variables were added in the subsequent model (Supplementary Table S4). The inclusion of sample-level variables showed the most substantial improvement (model 2 vs model 3: χ² (df = 5) = 9354·2, p <0·001), followed by the inclusion of time of day (χ² (df = 23) = 2511·1, p < 0·001). Therefore, model 4, incorporating patient-level, sample-level, and time of day variables, was selected as the final model. Blood glucose levels were significantly affected by time of day (p<0·001), with a peak of 9·6 [9·5 – 9·6; estimated marginal means, 95% CI] mmol/L at 10:00 in the morning and a trough of 8·6 [8·5 – 8·6] mmol/L at 03:00 at night (Table 2; Figure 3A). The effect of diabetes on glucose levels was found to be most pronounced (2·2 [2·1 – 2·3; coefficient, 95% CI], p<0·001), followed by an administration rate of carbohydrates > 8.5 grams/hour (1·3 [1·3 – 1·4], p<0·001) and administration of glucocorticoids (1·1 [1·0 – 1·1], p<0·001) (Table 2). Model diagnostics showed no violation of model assumptions (Supplementary Figure S4).

**Table 2:** Summary of final linear mixed-effects model.

| **Fixed effects** |  |  |  |
| --- | --- | --- | --- |
| (Intercept) | 6.6 | 6.5 – 6.7 | <0.001 |
| Time of day 03:00 [ref = 00:00]<sup>a</sup> | -0.4 | -0.5 – -0.4 | <0.001 |
| Time of day 10:00 [ref = 00:00]<sup>a</sup> | 0.6 | 0.5 – 0.7 | <0.001 |
| Age 56-65 years [ref = <55 years] | 0.5 | 0.3 – 0.6 | <0.001 |
| Age 66-75 years [ref = <55 years] | 0.6 | 0.5 – 0.7 | <0.001 |
| Age >75 years [ref = < 5 years] | 0.6 | 0.5 – 0.7 | <0.001 |
| Male sex [ref = Female] | -0.2 | -0.3 – -0.1 | <0.001 |
| Diabetes diagnosis [ref = No] | 2.2 | 2.1 – 2.3 | <0.001 |
| Carbohydrate administration 4.5<x≤6.5 grams/hour [ref = ≤4.5 grams/hour] | 0.6 | 0.6 – 0.7 | <0.001 |
| Carbohydrate administration 6.5<x≤8.5 grams/hour [ref = ≤4.5 grams/hour] | 0.9 | 0.9 – 0.9 | <0.001 |
| Carbohydrate administration >8.5 grams/hour [ref = ≤4.5 grams/hour] | 1.3 | 1.3 – 1.4 | <0.001 |
| Insulin administration 0<x≤200 units/hour [ref = 0 units/hour ] | -0.5 | -0.5 – -0.4 | <0.001 |
| Insulin administration 200<x≤750 units/hour [ref = 0 units/hour ] | 0.9 | 0.9 – 0.9 | <0.001 |
| Insulin administration >750 units/hour [ref = 0 units/hour ] | 0.7 | 0.6 – 0.7 | <0.001 |
| Dextrose administration 0<x≤0.5 grams/hour [ref = 0 grams/hour ] | 0.1 | 0.0 – 0.1 | 0.002 |
| Dextrose administration 0.5<x≤2 grams/hour [ref = 0 grams/hour ] | 0.2 | 0.2 – 0.2 | <0.001 |
| Dextrose administration >2 grams/hour [ref = 0 grams/hour ] | 0.6 | 0.6 – 0.7 | <0.001 |
| Glucocorticoid administration [ref = No] | 1.0 | 1.0 – 1.1 | <0.001 |
| **Variance** |  |  |  |
| Inter-patient random effect (intercept) | 2.8 |  |  |
| Residual | 5.4 |  |  |

<sup>a</sup> Time of day was included as a categorical variable with 24 levels of 1-h time bins; only 03:00 (trough) and 10:00 (peak) are shown here due to space constraints. Estimates at all time bins are shown in Figure 3A.
<sup>b</sup> The start and end time of administration of insulin and glucocorticoids (end times only) were corrected to account for the delaying and/or persisting effect of insulin and glucocorticoids on glucose levels (see Methods for details).
CI: confidence interval; ref = reference

**Figure 3:**
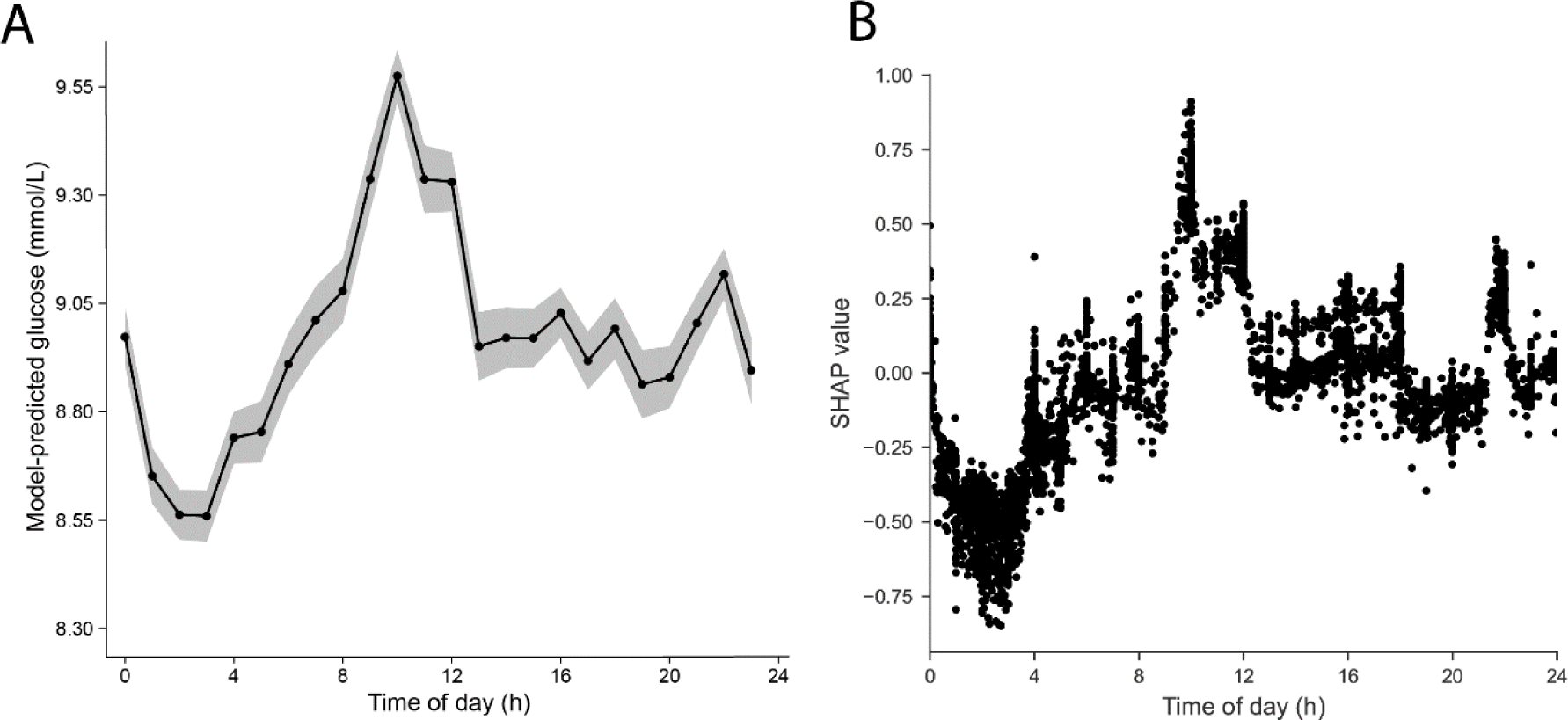
24-h variation in glucose levels during the entire ICU stay during enteral nutrition. (**A**) Model-predicted glucose levels by time of day (in hourly bins). Data presented as estimated marginal means ± 95% CI derived from the final linear mixed effects model, adjusted for patient-level and sample-level covariates. (**B**) SHAP values for time of day (as a continuous variable) from the XGBoost model. Each black dot represents an individual glucose measurement. The SHAP value represents shows how much the time of day feature affected the glucose level. A positive SHAP value reflects a positive impact on the predicted glucose level, while negative values reflect negative impacts. Its absolute value represents the magnitude of this effect. A sample of 5000 randomly selected features of the glucose measurements was used in this SHAP analysis.

Following selection of the optimal XGBoost model by hyperparameter tuning (optimal parameters: maximum tree depth: 5; Learning rate: 0·1; Number of estimators: 150; Minimum child weight: 10; Fraction of columns to be subsampled in each tree: 0·75; Subsample ratio of training instances: 0·75), a 24-hour pattern was observed in the SHAP values for time of day, with an increase around 10:00h and a decrease between 01:00h and 03:00h (Figure 3B), resembling the 24-hour pattern observed with the linear-mixed effects model (Figure 3A). Time of day was the fifth most important feature, preceded by diabetes, carbohydrate and insulin administration rate and glucocorticoid administration, and followed by age, dextrose administration and sex (Supplementary Figure S5A-B). SHAP dependence plots for continuous features (carbohydrate, insulin and dextrose administration rate and age) are shown in Supplementary Figure S6A-D, in which the relationship between values of features and SHAP values can be assessed.

In all subgroups of type of mechanical ventilation, in-hospital mortality, RASS-scores, sample type, days in ICU and sampling frequencies, similar 24-hour patterns were observed, with expected differences in 24-hour baseline levels (Figure 4). Remarkably, the peak-to-trough difference in glucose levels was more than two-fold larger in the subgroup of patients who received > 70 units of insulin per day during their stay (9·6 [9·4 – 9·9] versus 10·7 [10·4 – 10·9] mmol/L at 03:00 and 10:00, respectively) and 35-75 units of insulin per day (10·3 [10·0 – 10·6] versus 11·8 [11·6 – 12·1] at 03:00 and 10:00, respectively), compared to the subgroup of patients who did not require insulin therapy (7·0 [7·0 – 7·1] mmol/L at 03:00 and 7·5 [7·4 – 7·6] at 10:00) (Figure 5).

**Figure 4:**
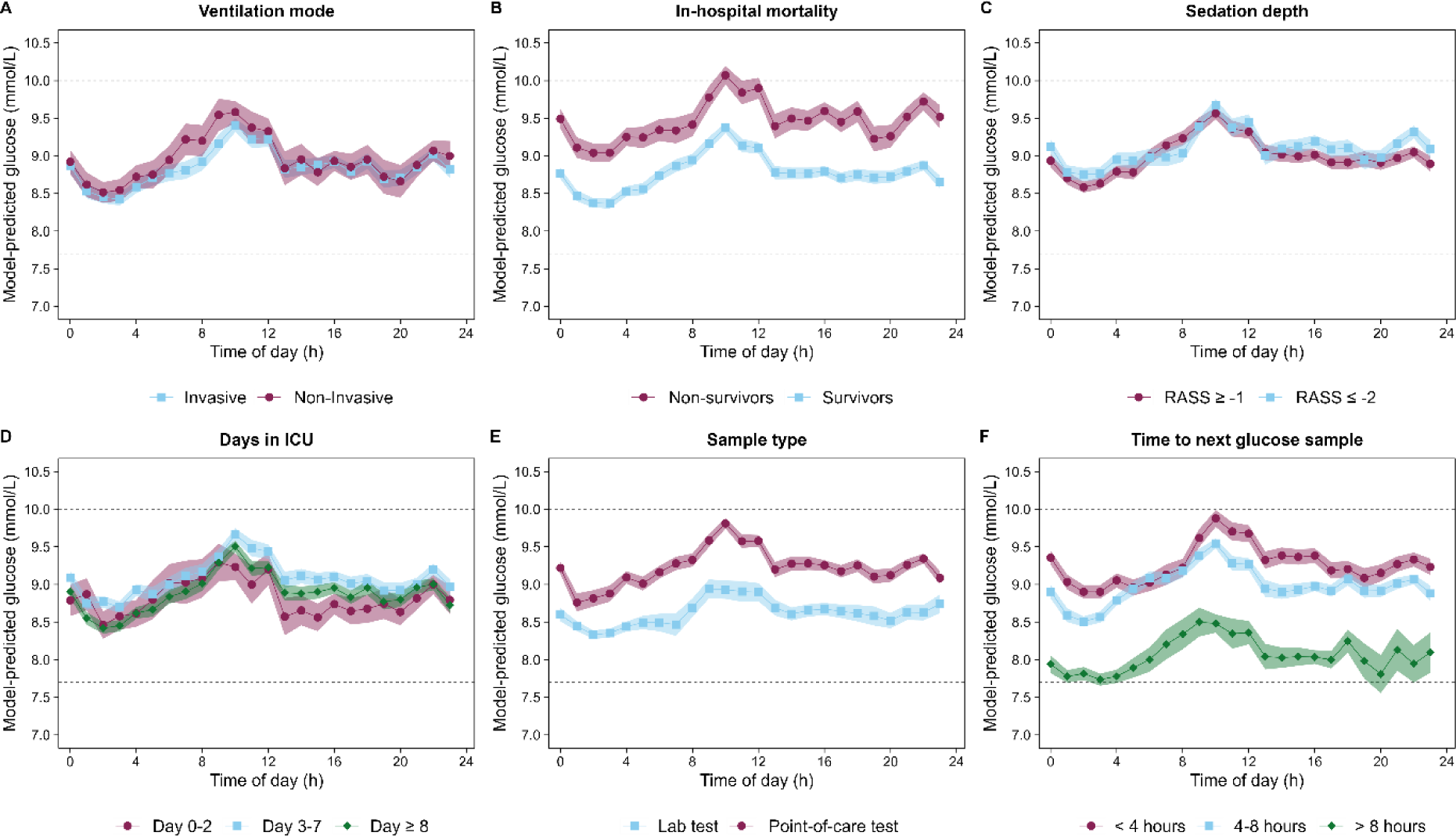
Subgroup analysis of 24-hour variation in glucose levels. (**A**) Ventilation mode. (**B**) Survivor status. (**C**) Depth of sedation. (**D**) Days in ICU. (**E**) Sample type. (**F**) Time to next glucose sample. For each subgroup, a linear mixed-effects model was fitted. Data presented as estimated marginal means ± 95% CI, adjusted for covariates. Number of patients and measurements per group are shown in Supplementary Table S5.

**Figure 5:**
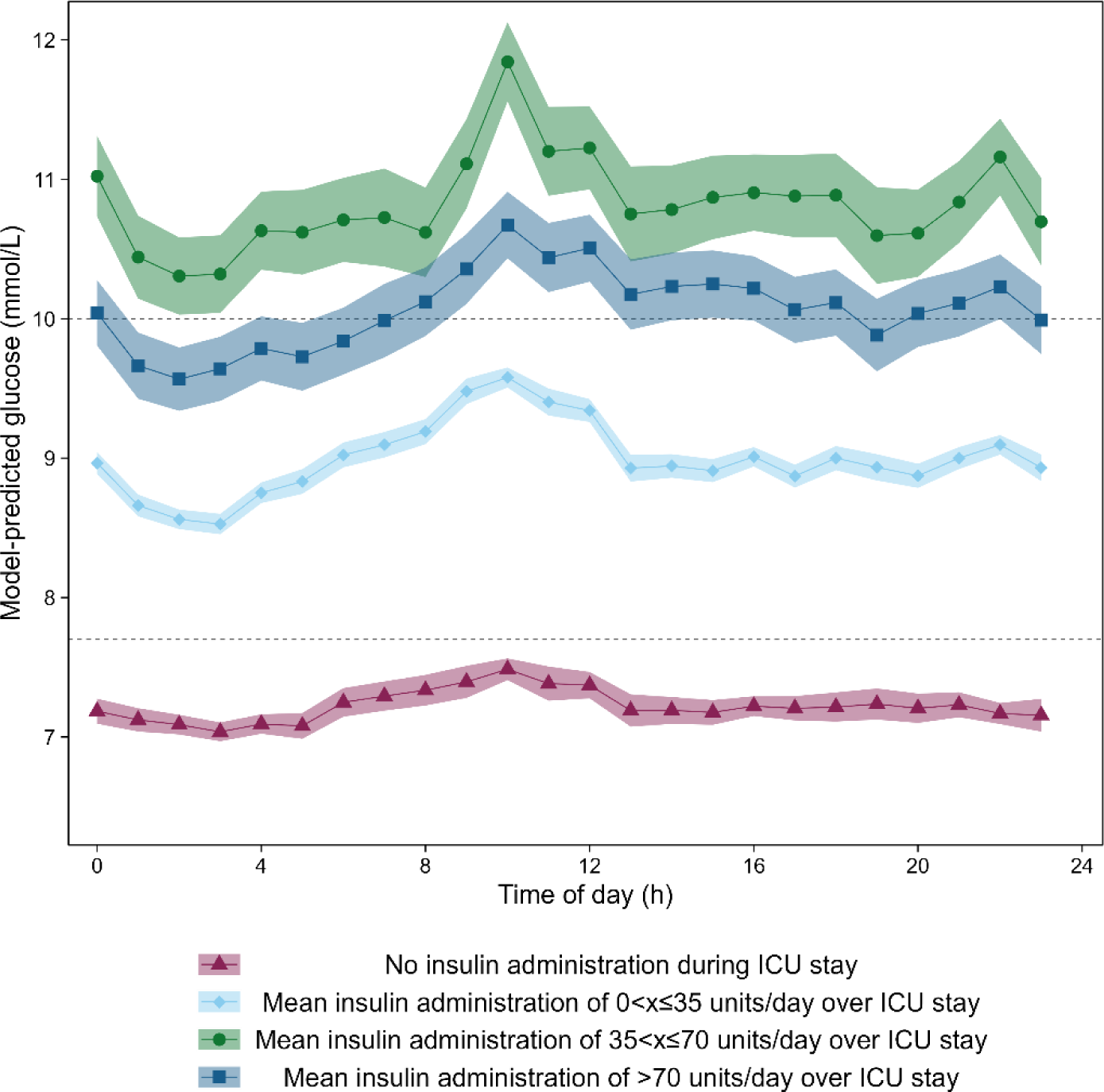
24-h variation in glucose levels in patient who required insulin during their stay compared to those who did not receive any insulin. Data presented as estimated marginal means ± 95% CI, adjusted for covariates.

## Discussion

Our results indicate that glucose levels in critically ill patients show marked 24-hour variation during continuous enteral nutrition. This daily pattern persists when controlling for potential sources of bias and confounding factors, suggesting that the observed 24-hour variation is due to biological processes rather than health care processes, patient characteristics, or treatment effects. These findings were confirmed with an alternative machine learning approach, thereby showing the consistency of the results obtained with the linear mixed-effects model. Remarkably, the effect of time of day on glucose levels, even during continuous nutritional support, is equal to or even larger than covariates that are commonly considered in analyses, such as sex and age. This highlights the importance of incorporating time of day in analyses of biological time series data.

We found a decrease of glucose levels during nighttime, followed by a substantial rise of glucose levels in the early morning. Previous studies also observed a nocturnal decrease in glucose levels in critically ill patients, although the timing of the trough varied among studies.^9–11^ However, those studies included glucose measurements regardless of nutritional status or administration of medication of fluids that directly affect glucose levels, such as insulin, glucocorticoids, or dextrose. For example, the daily variation in glucose levels observed in previous studies may therefore be driven by an interfering daily pattern in nutrition, particularly when considering oral intake. Since most oral food intake presumably occurs during the daytime and not during the nighttime, any observed 24-hour pattern in glucose levels may be attributable to patterns in food intake rather than endogenous variation in glucose control. A strength of our study is that we only included glucose measurements that were taken while patients received enteral nutrition in order to investigate 24-hour variation in glucose levels during nutritional intake. To verify whether the observed daily variation in glucose levels is driven by patients who have (daytime) oral intake (which may not be well documented in the EHR), we performed subgroup analyses on data from sedated patients (with RASS score ≤ -2) and patients on invasive mechanical, who can be presumed not to have any oral food intake. These analyses show that the 24-hour pattern persists in these subgroups, ruling out an effect of oral food intake. In addition, we verified that the timing of glucose measurements, enteral nutrition, and administration of insulin, dextrose, and glucocorticoids were equally distributed over the 24-hour period and included these as covariates in the model.

In addition to the subgroup analyses to control for patients with potential oral intake, we performed further subgroup analyses to assess the effect of other potential sources of bias that were not or insufficiently available in the MIMIC-IV database. Subgroups were created based on available variables that might indirectly affect glucose levels. Firstly, the glucose sampling method used (lab or point-of-care tests) can be influenced by health care processes and acuteness of situation. Lab tests are more likely to be a routine test, collected as part of a panel of tests, while point-of-care tests can be more likely to performed when glucose needs to be closely monitored. Besides, the sampling method used could have affected the measured glucose levels, although this effect was shown limited in the MIMIC-III database.^19^ Also, severity of illness could affect the glucose levels. Indicators of severity of illness, such as Sequential Organ Failure Assessment (SOFA) scores, were not consistently present in the data. Therefore, in-hospital mortality and days in the ICU were regarded to be associated with illness severity. To assess the effect of sampling bias, time to the next glucose measurements was used a proxy of sampling frequency. For all these potential sources of bias, the expected differences in mean 24-hour levels were observed, but the 24-hour pattern with the morning rise and its timing remained consistent among the subgroups.

Linear mixed-effects models prove to be valuable in the analyses conducted in our study as this approach allows for adjustment of several factors with a known influence on the dependent variable (fixed effects) and accounts for individual variability (random effects). As an addition to the main analyses, the same dataset was analyzed with an XGBoost regression model. XGBoost is a widely adopted machine learning algorithm that uses decision trees within a gradient boosting framework. Although XGBoost is commonly employed for prediction tasks, it is used in our study to assess the effect of a variable of interest (time of day) on an observed parameter (glucose levels), as was also done previously.^26,27^ While linear mixed-effects models assume a linear relationship between fixed effects and outcome variable, XGBoost can capture more intricate patterns and non-linear associations that might exist in the data. This allowed us to incorporate age and administration rates of carbohydrates, insulin and dextrose as continuous variables in the XGBoost models. Consequently, we revealed the pattern of the non-linear relationship between insulin administration rate and glucose levels. However, a notable drawback of the XGBoost model is its inability to account for inter-patient variability and repeated measurements, as is possible with the random effects in the linear mixed-effects model. In the end, similar results were found when analyzing the data with an XGBoost model as with the linear mixed-effects model, which shows consistency of our findings. Moreover, this indicates that the use of a machine learning model may be a powerful alternative to traditional linear mixed-effects models in the analysis of time series data.

The 24-hour pattern in glucose levels with low levels at night and an increase in the early morning that we observe in our study is markedly different from what could be expected based on studies on circadian rhythms in glucose control in healthy individuals. In healthy individuals, a complex interplay of peripheral circadian clocks in different tissues regulate the physiological processes involved in glucose metabolism in a time of day-dependent manner, resulting, for example, in circadian variation in circulating ghrelin^28^ and leptin levels^29^, pancreatic insulin secretion^30^, and whole-body insulin sensitivity^31^. In general, this results in higher glucose excursion when food is consumed at the wrong time of day, i.e. during the biological night compared to the day^25^. A study among healthy male participants who received continuous enteral nutrition in controlled laboratory conditions revealed that glucose levels are highest during the second half of the night and lowest during the day, i.e., a 24-hour rhythm that is advanced by several hours compared to the pattern observed in our study.^32^ However, the sharp rise in blood glucose levels observed in our analysis during the early morning is reminiscent of the dawn phenomenon observed in people with diabetes mellitus. This phenomenon occurs as a result of reduced insulin sensitivity at the time of highest endogenous glucose production.^3^ The morning increase that we observe in critically ill patients may reflect increased insulin resistance that is frequently observed in the ICU population.^33^ This notion gains further support from the observation that patients who require insulin – who are thus likely to have impaired glucose tolerance, or even a certain degree of insulin resistance – exhibited a more pronounced morning peak compared to patients who did not require insulin. Albeit correlational, our findings add real-world support to the body of literature from pre-clinical studies and studies in healthy human participants that show that glucose metabolism is anything but constant across the 24-hour period.^2^ The exact mechanisms that underlie the observed pattern, e.g., 24-hour variation in local insulin sensitivity, pancreatic β-cell function, fever and/or circulating levels of other hormones involved in glucose metabolism (e.g., cortisol, leptin, or ghrelin) remains to be uncovered in future (experimental) studies.

Monitoring of blood glucose levels is an essential aspect of critical care management, as critically ill patients are prone to high glycemic variations. Our study contributes to the understanding of daily fluctuations of glucose levels in patients during clinical practice at the ICU. With this understanding, glucose levels can be interpreted in the context of the time of day. Nonetheless, we recognize that the variation of about 1 mmol/L across 24 hours may not affect glucose monitoring or insulin administration protocols in the ICU. Also, the substantial effect of time of day on blood glucose levels observed in our study emphasizes the importance of considering the 24-hour variation in scientific research on glucose regulation in critically ill patients. Relying solely on single measurements or averages derived from unequally distributed measurements may inaccurately reflect the average daily glucose control.^13^ Likewise, clinical trials investigating glucose control in critically ill patients may need to consider this daily variation in their sampling protocol and ensuing study endpoints.

One of the strengths of our study is the large number of glucose measurements that was included in the analysis. The availability of a large, rich, clinical dataset enabled the adjustment of multiple confounding factors. Additionally, a heterogeneous group of patients admitted to the ICU was included, thereby making the results more applicable to the mixed ICU population. Nonetheless, several limitations should be considered when interpreting our findings. The retrospective design introduces inherent limitations and a certain degree of uncontrolled variability.^34^ As a result, it was not possible to control the timing of blood glucose measurements or the ensuing medical decisions. Being constrained to data available in the electronic health records, we had to deal with a limited number of samples per patient per day and with the potential of incomplete data. Consequently, a limitation is that we assumed that the lack of recorded events, diabetes ICD-codes, or date of death data within the MIMIC-IV dataset indicated their absence in real-life. Unfortunately, we were unable to extract information about glucocorticoid dose and therefore had to restrict our analysis to a binary classification on whether or not patients received glucocorticoids, precluding us from evaluating its impact on glucose levels in more detail. In the end, with the chosen variables, all data required to construct the variables was available for each glucose measurement. Another limitation is introduced by the variability in the delay and persisting effects after the start and end of administration of the assessed medication on glucose levels. We incorporated fixed correction times to the start and end times of insulin and glucocorticoid administration to account for their duration of action, but any interindividual variability in the duration or magnitude of their effect was not taken into account. Also, the use of single-center data in this study may introduce bias due to center-specific factors, patient population, or variations in patient management practices. Moreover, it remains to be investigated to what extent patient-level factors that influence 24-hour patterns in glucose levels impact our findings, such as sleep^35–37^, neurotrauma^38^, or circadian clock function.^39^ In general, our study provides insight into the glucose control during continuous enteral nutrition in critically ill patients. An interesting question is to what extent the administration of continuous enteral nutrition gives rise to the 24-hour pattern in blood glucose values and is responsible for the high prevalence of insulin resistance that is observed in ICU patients in general. Although this type of nutritional support is common practice in ICUs worldwide, it is known from pre-clinical work and studies in healthy human participants that food intake at the wrong time of day (i.e. during the biological night) leads to unfavorable glucose responses.^2^ Indeed, it has been questioned whether continuous nutrition is the most optimal nutrition schedule from various perspectives (incl. glycemic control, circadian rhythmicity, muscle metabolism, and gastrointestinal function).^40,41^ Future studies that compare the effect of continuous feeding to other schedules on various clinical outcomes could additionally investigate the impact on daily glucose variability.

To conclude, our findings demonstrate the presence of a substantial 24-hour pattern in blood glucose levels during continuous enteral nutrition in critically ill patients that persists after adjustment for relevant sources of bias. This suggests that the observed 24-hour variation is due to the endogenous biological variation rather than as a result of bias introduced by health care processes, patient characteristics, or treatment effects. In addition, our study provides a framework for addressing these sources of bias when analyzing the effect of time of day on physiological variables available in electronic health records. Our findings contribute to a deeper understanding of daily fluctuations in glucose levels observed at the bedside by nurses and clinicians working at the ICU. Future research should focus on multicenter cohorts and incorporate individual circadian phase and brain states in the analysis to further enhance our understanding of these observations.

## Data Availability

The MIMIC-IV dataset is publicly available via: https://mimic.mit.edu/. The code used for the curation of the dataset and analyses are available at https://github.com/fwhiemstra/mimic-glucose-24h-variation-in-icu.

https://mimic.mit.edu/

## Contributors

The study was conceptualized by LK. Data extraction, preprocessing and analysis was performed by FWH and LK. DJvW, EdJ, DJS and AK participated in discussing and interpretation of the results. FWH wrote the first version of the manuscript. All authors have edited and reviewed the manuscript and approved the final version.

## Declaration of interest

All authors declare no competing interests.

## Acknowledgements

The authors thank Marc D. Ruben for valuable discussions. This work was supported by a VENI grant (2020-09150161910128 to L.K.) from the Netherlands Organization for Health Research and Development (ZonMw), an institutional project grant from the Leiden University Medical Center (to L.K. and D.J.v.W.) and the BioClock Consortium (project number 1292.19.077 to LK) funded by the research programme NWA-ORC by the Dutch Research Council (NWO). We would like to thank the researchers of the MIT Computational Physiology Laboratory and collaborating researchers for developing, maintaining and publicly sharing of the MIMIC-IV clinical database.

## Notes

### Competing Interest Statement

The authors have declared no competing interest.

### Author Declarations

The Institutional Review Board at the Beth Israel Deaconess Medical Center granted a waiver of informed consent for the data collected in the MIMIC-IV database (that was used in this study).

### Summary of Updates

In this revised manuscript, we now quantified administration of dextrose (grams/hours), insulin (units/hour) and carbohydrate (grams/hour) in nutrition (grams/hour), which provides more granularity to our analysis and insight into our findings. The observed 24-hour pattern remained similar to the results of our previous analysis. In addition, we performed more detailed analysis of patients without and with insulin requirement (who are thus considered to have impaired glucose tolerance, or even a certain degree of insulin resistance), showing that the peak-to-trough excursion is roughly twice as high in patients who required insulin therapy during their ICU stay - and are thus considered insulin resistant - compared to those who are not. Lastly, we elaborated more on the limitations of our analysis, including the retrospective use of electronic health record data and thereby being constrained to the data available.

