## Supplementary material for "Daily variation in blood glucose levels during continuous enteral nutrition in patients on the Intensive Care Unit: a retrospective observational study"

### TABLE OF CONTENTS

**SUPPLEMENTARY METHODS** ........ 3

Supplementary Methods S1: XGBoost analysis using SHAP values
Supplementary Methods S2: Formulas of linear mixed-effects models

**SUPPLEMENTARY TABLES** ........ 4

Supplementary Table S1: Carbohydrate content for each nutrition product present in the dataset.
Supplementary Table S2: Overview of variables included in the linear mixed-effects models and XGBoost model.
Supplementary Table S3: Characteristics of included glucose measuring (during enteral feeding episodes).
Supplementary Table S4: Results of log-likelihood ratio test to compare model fits.
Supplementary Table S5: Number of glucose measurements and patients for each subgroup in sensitivity analysis.

**SUPPLEMENTARY FIGURES** ........ 7

Supplementary Figure S1: Overview of variables included in the statistical analysis.
Supplementary Figure S2: Proportion of glucose measurements taken during administration of insulin, dextrose, and glucocorticoids by time of day.
Supplementary Figure S3: Glucose levels over time, normalized per patient.
Supplementary Figure S4: Model diagnostic plots of final linear mixed-effects model.
Supplementary Figure S5: SHAP analysis of XGBoost regression model.
Supplementary Figure S6: SHAP dependence plots of carbohydrate administration rate, insulin administration rate, dextrose administration rate and age.

**REFERENCES** ........ 13

#### SUPPLEMENTARY METHODS

##### Supplementary Methods S1: XGBoost analysis using SHAP values

In order to show robustness of our results obtained with the linear mixed-effects model, the analysis was repeated using the same dataset with an Extreme Gradient Boosting (XGBoost) regression model.[1] XGBoost is a widely adopted machine learning algorithm that uses ensembling of decision trees within a gradient boosting framework and is known for its computational efficiency and good performance. XGBoost analysis and visualization was performed using Python programming language (version 3.10.2) using xgboost package version 1.7.6[1], shap package version 0.41.0[2] and scikit-learn version 1.3.0.[3]

The variables that were used as fixed effects in linear mixed-effects model 4 were used as features in a XGBoost regression model: age, sex, diabetes diagnosis, time of day, administration rates of carbohydrate (grams/hour), dextrose (grams/hour) and insulin (units/hour) and administration of glucocorticoids (yes/no). In contrast to linear mixed-effects model, time of day was used as continuous variable (in decimal units from 0 to 24) in the XGBoost regression model (ranging from 0 to 24), and age and administration rates of carbohydrate, insulin and dextrose were used as continuous variables. Glucose level (in mmol/L) was the target variable (Supplementary Table S1).

Tuning of hyperparameter (i.e. a parameter that is set before the learning process and whose value is used to control the learning process) is essential to optimize model performance and generalization. We used a grid search with 5-fold cross-validation for hyperparameter tuning. The ranges of hyperparameters for the grid search were as follows: 1) Maximum tree depth: 5, 10, 15, 25, 50; 2) Learning rate: 0·05, 0·1, 0·3; 3) Number of estimators: 25, 50, 100, 150; 4) Minimum child weight: 1, 5, 10; 5) Fraction of columns to be subsampled in each tree; 0·75, 1·0; 6) Subsample ratio of training instances: 0·75, 1·0. Default settings of the xgboost Python package were used for the rest of the hyperparameters.[1]

In order to assess the effect of time of day (and the other variables included as features in the model) on glucose levels, the SHapley Additive exPlanations (SHAP) method was used.[2] SHAP is a method used in machine learning to explain how each feature contributes to the model's prediction. It assigns importance scores (SHAP values) to each feature for each single observation (which is in our study a glucose measurement) when considered in combination with all features in the model. The magnitude of the SHAP value thus represents the increase or decrease in the glucose level that can be attributed to the feature value.

##### Supplementary Methods S2: Formulas of linear mixed-effects models

| | |
|---|---|
| Model 1 | `glucose_level ~ 1 + (1 \| subjectID)` |
| Model 2 | `glucose_level ~ diabetes + age + sex + (1 \| subjectID)` |
| Model 3 | `glucose_level ~ diabetes + age + sex + carbohydrate_administration_rate + insulin_administration_rate + insulin_administration_rate + glucocorticoid_administration + (1 \| subjectID)` |
| **Model 4** | `glucose_level ~ diabetes + age + sex + carbohydrate_administration_rate + insulin_administration_rate + insulin_administration_rate + glucocorticoid_administration + time_of_day (1 \| subjectID)` |

### SUPPLEMENTARY TABLES

#### Supplementary Table S1: Carbohydrate content for each nutrition product present in the dataset.

| Nutrition type | Carbohydrate content | Source |
|---|---|---|
| **Beneprotein** | 0 | https://www.nestlehealthscience.ca/en/brands/beneprotein/beneprotein-hcp |
| **Boost Glucose Control** | 0.068 | https://www.boost.com/products/glucose-control |
| **Enlive** | 0.190 | Fitgerald et al. (2021)[4] |
| **Ensure** | 0.139 | Fitgerald et al. (2021)[4] |
| **Ensure Plus** | 0.203 | Fitgerald et al. (2021)[4] |
| **Fiber Supplement (i.e. Banana Flakes)** | NOT FOUND | N/A |
| **Fibersource HN** | 0.156 | Fitgerald et al. (2021)[4] |
| **Glucerna** | 0.11 | Fitgerald et al. (2021)[4] |
| **Impact** | 0.132 | Fitgerald et al. (2021)[4] |
| **Impact with Fiber** | 0.132 | Fitgerald et al. (2021)[4] |
| **Isosource 1.5** | 0.167 | Fitgerald et al. (2021)[4] |
| **Jevity 1.2** | 0.170 | https://www.abbottnutrition.com/our-products/jevity-1_2-cal |
| **Jevity 1.5** | 0.216 | https://www.abbottnutrition.com/our-products/jevity-1_5-cal |
| **Nepro** | 0.147 | Fitgerald et al. (2021)[4] |
| **NovaSource Renal** | 0.185 | Fitgerald et al. (2021)[4] |
| **Nutren 2.0** | 0.196 | Fitgerald et al. (2021)[4] |
| **Nutren Pulmonary** | 0.10 | Fitgerald et al. (2021)[4] |
| **Nutren Renal** | 0.185 | Fitgerald et al. (2021)[4] |
| **Osmolite 1.5** | 0.203 | Fitgerald et al. (2021)[4] |
| **Peptamen 1.5** | 0.188 | Fitgerald et al. (2021)[4] |
| **Peptamen Bariatric** | 0.078 | Fitgerald et al. (2021)[4] |
| **ProBalance** | NOT FOUND | N/A |
| **Promote** | 0.131 | https://www.abbottnutrition.com/our-products/promote |
| **Promote with Fiber** | 0.139 | https://www.abbottnutrition.com/our-products/promote-with-fiber |
| **Pulmocare** | 0.105 | Fitgerald et al. (2021)[4] |
| **Replete** | 0.112 | Fitgerald et al. (2021)[4] |
| **Replete with Fiber** | 0.124 | Fitgerald et al. (2021)[4] |
| **Two Cal HN** | 0.219 | Fitgerald et al. (2021)[4] |
| **Vital 1.5** | 0.186 | https://www.abbottnutrition.com/our-products/vital-1_5-cal |
| **Vital High Protein** | 0.113 | https://www.abbottnutrition.com/our-products/vital-hp |
| **Vivonex** | 0.003 | Fitgerald et al. (2021)[4] |

#### Supplementary Table S2: Overview of variables included in the linear mixed-effects models and XGBoost model.

| | Linear mixed-effects model | | | | | | XGBoost model | |
|---|---|---|---|---|---|---|---|---|
| | **Variable type** | **Variable in model** | **Model 1** | **Model 2** | **Model 3** | **Model 4** | **Variable type** | **Variable in model** |
| **Glucose level** | Continuous | Dependent variable | X | X | X | X | Continuous | Target variable |
| **PatientID** | Categorical | Random effect | X | X | X | X | | |
| **Intercept** | | | X | X | X | X | | |
| **Patient level variables** | | | | | | | | |
| **Age (years)** | Categorical [≤55, 55<x≤65, 65<x≤75 >75] | Fixed effect | | X | X | X | Continuous | Feature |
| **Sex** | Categorical [female / male] | Fixed effect | | X | X | X | Categorical [female, male] | Feature |
| **Diabetes diagnosis** | Categorical [yes, no] | Fixed effect | | X | X | X | Categorical [yes, no] | Feature |
| **Sample level variables** | | | | | | | | |
| **Carbohydrate administration rate (grams/hour)** | Categorical [≤4.5, 4.5<x≤6.5, 6.5<x≤8.5 >8.5] | Fixed effect | | | X | X | Continuous | Feature |
| **Insulin administration rate (units/hour)[a]** | Categorical [0, 0<x≤200, 200<x≤750, >750] | Fixed effect | | | X | X | Continuous | Feature |
| **Dextrose administration rate (grams/hour)[a]** | Categorical [0, 0<x≤0.5, 0.5<x≤2, >2] | Fixed effect | | | X | X | Continuous | Feature |
| **Glucocorticoid administration[a]** | Categorical [yes, no] | Fixed effect | | | X | X | Categorical [yes, no] | Feature |
| **Time variable** | | | | | | | | |
| **Time of day** | Categorical [time bins 0 to 23] | Fixed effect | | | | X | Continuous | Feature |

[a] The start and/or end time of administration of insulin, dextrose and glucocorticoids were corrected to account for their delaying and/or persisting effect on glucose levels (see Methods for details).

#### Supplementary Table S3: Characteristics of included glucose measuring (during enteral feeding episodes).

| Glucose measurement characteristics | Included glucose measurements (n=207,647) |
|---|---|
| **Glucose measurements per patient per day, median [IQR]** | 3 [2-5] |
| **Glucose measurements taken during administration of:, n (%)** | |
| **Dextrose[a]** | 89,581 (43%) |
| **Insulin[a]** | 70,066 (34%) |
| **Glucocorticoids[a]** | 40,314 (19%) |
| **Sample type, n (%)** | |
| **Finger stick** | 129,133 (62%) |
| **Lab (whole blood)** | 64,489 (31%) |
| **Lab (serum)** | 14,025 (6.8%) |

[a] The start and/or end time of administration of insulin, dextrose and glucocorticoids were corrected to account for their delaying and/or persisting effect on glucose levels (see Methods for details).

#### Supplementary Table S4: Results of log-likelihood ratio test to compare model fits.

| | Number of variables | Log-Likelihood | Degrees of freedom | Chi-squared value | P-value |
|---|---|---|---|---|---|
| **Model 1** | 3 | -484169 | | | |
| **Model 2** | 8 | -483164 | Compared to model 1 | | |
| | | | 5 | 2009.5 | <0.0001 |
| **Model 3** | 18 | -478487 | Compared to model 2 | | |
| | | | 10 | 9354.2 | <0.0001 |
| **Model 4** | 41 | -477232 | Compared to model 3 | | |
| | | | 23 | 2511.1 | <0.0001 |

#### Supplementary Table S5: Number of glucose measurements and patients for each subgroup in sensitivity analysis.

| Variable | Number of glucose measurements | Number of patients |
|---|---|---|
| **Ventilation mode** | | |
| **Non-Invasive** | 20,408 | 2,416 |
| **Invasive** | 78,715 | 4,587 |
| **Survivor status** | | |
| **Non-survivors** | 59,523 | 1,967 |
| **Survivors** | 148,124 | 4,962 |
| **Sedation depth** | | |
| **RASS ≥ -1** | 96,191 | 5,191 |
| **RASS ≤ -2** | 81,903 | 4,532 |
| **Day in the ICU** | | |
| **Day 0-2** | 14,714 | 3,251 |
| **Day 3-7** | 80,839 | 6,276 |
| **Day ≥ 8** | 112,094 | 3,718 |
| **Sample type** | | |
| **Point-of-care test** | 129,133 | 6,199 |
| **Lab test** | 78,514 | 6,779 |
| **Time to next glucose measurement** | | |
| **< 4 hours** | 91,845 | 5,689 |
| **4-8 hours** | 94,694 | 6,224 |
| **> 8 hours** | 18,108 | 4,621 |
| **Insulin requirement** | | |
| **No insulin requirement** | 30,315 | 2,509 |
| **Mean daily insulin administration of insulin:** | | |
| **0<x<=35 units** | 110,374 | 3,499 |
| **35<x<=70 units** | 24,692 | 460 |
| **>70 units** | 42,266 | 461 |

#### SUPPLEMENTARY FIGURES

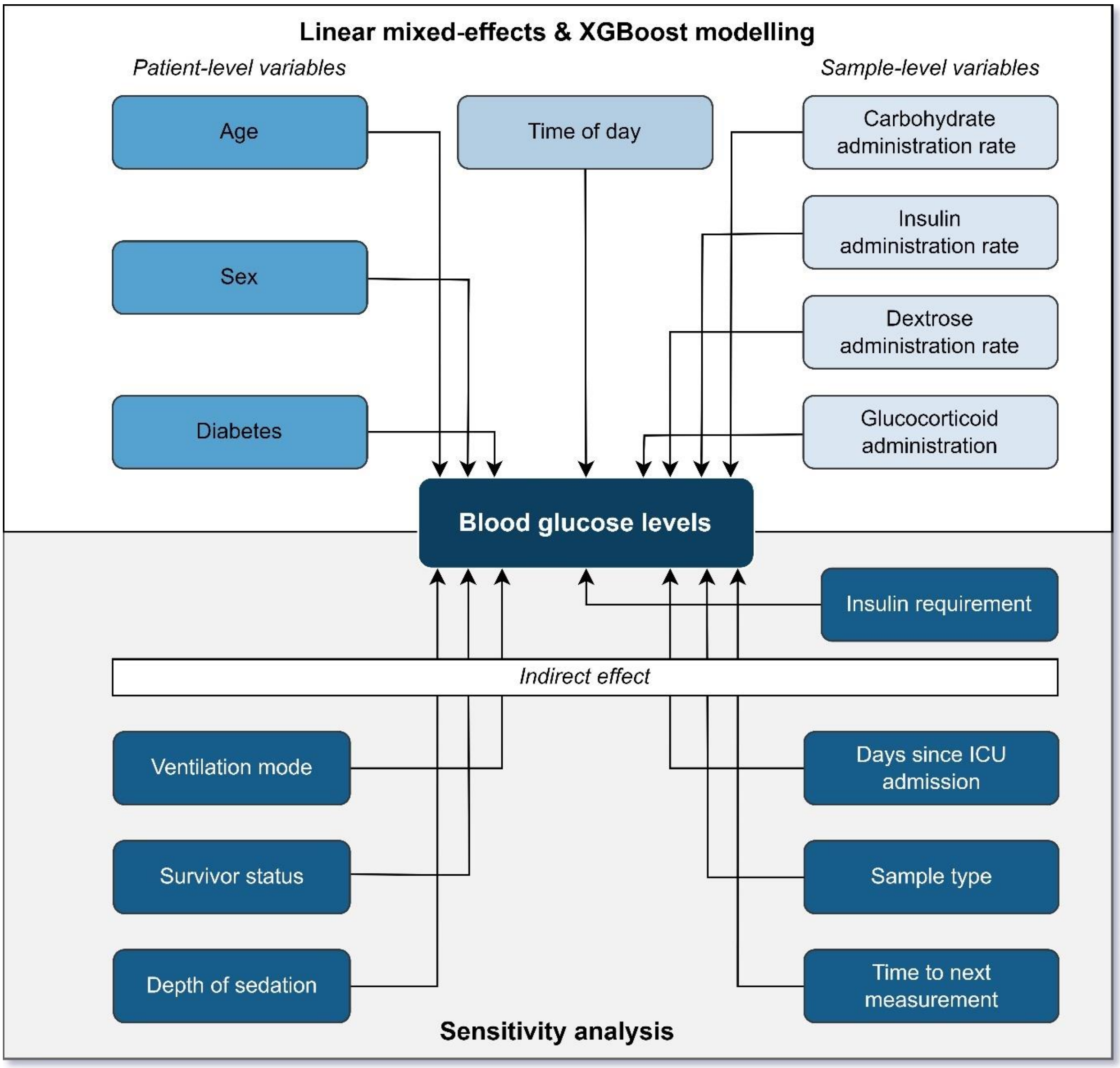


**Supplementary Figure S1: Overview of variables included in the statistical analysis.** Variables in the upper half of the figure are included in the linear mixed-effects models and XGBoost model. The variable groups (patient-level variables and sample-level variables) that are sequentially used in the various linear-mixed effects models (model 1-4) are represented in the figure. Variables in lower half of the figure are used to define the subgroups that are assessed in the sensitivity analyses.

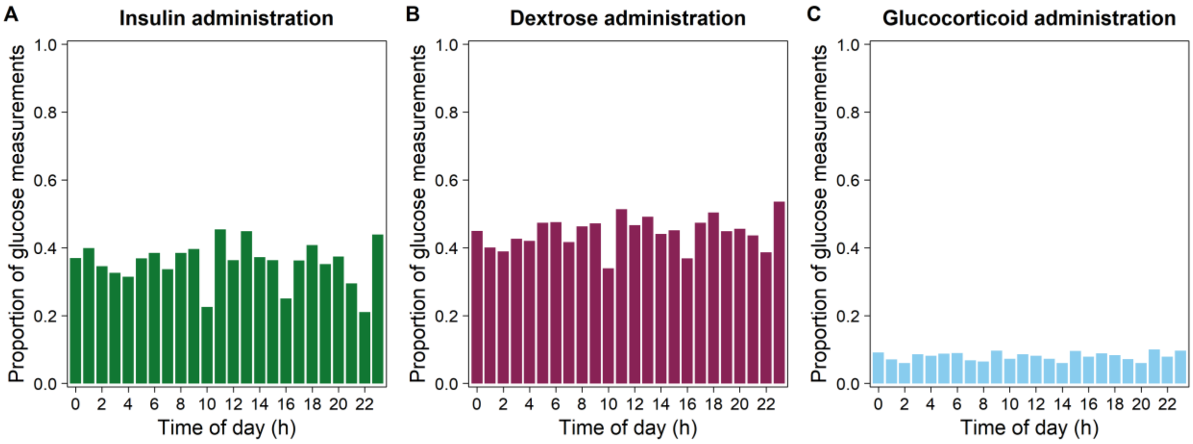


**Supplementary Figure S2: Proportion of glucose measurements taken during administration of (A) insulin, (B) dextrose, and (C) glucocorticoids by time of day.** The start and end time of administration of insulin and glucocorticoids (end times only) were corrected to account for the delaying and/or persisting effect of insulin and glucocorticoids on glucose levels (see Methods for details).

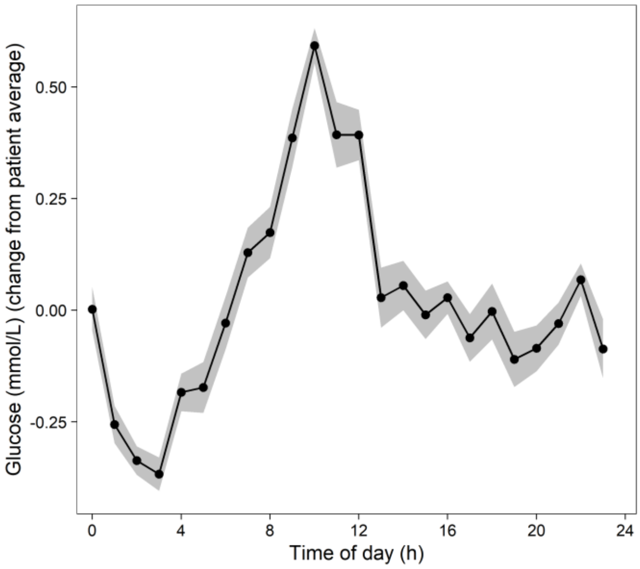

**Supplementary Figure S3: Glucose levels over time, normalized per patient.**

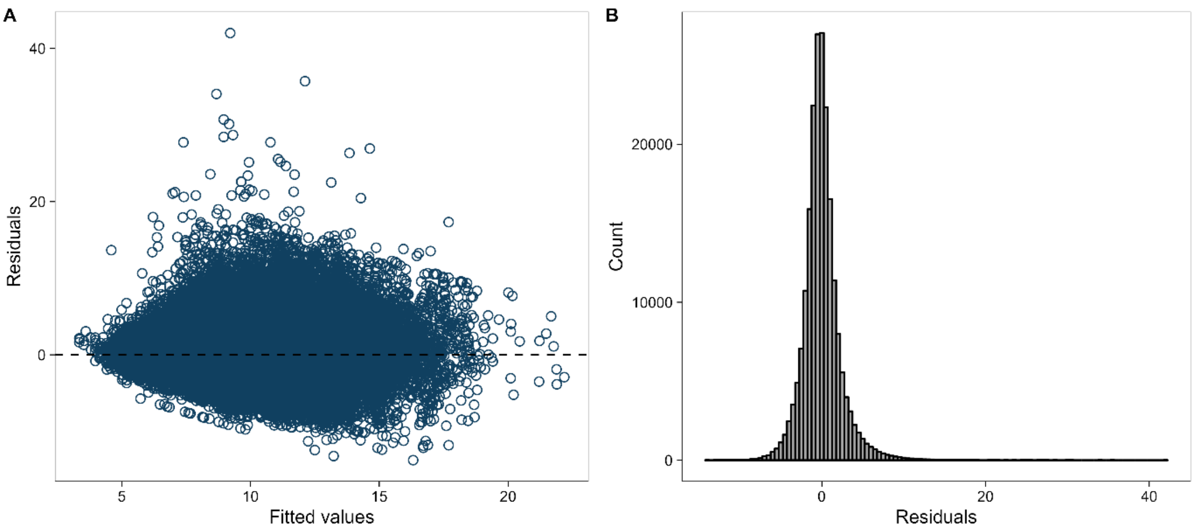


**Supplementary Figure S4: Model diagnostic plots of final linear mixed-effects model. (**A) Residuals versus fitted values. (B) Distribution of residuals.

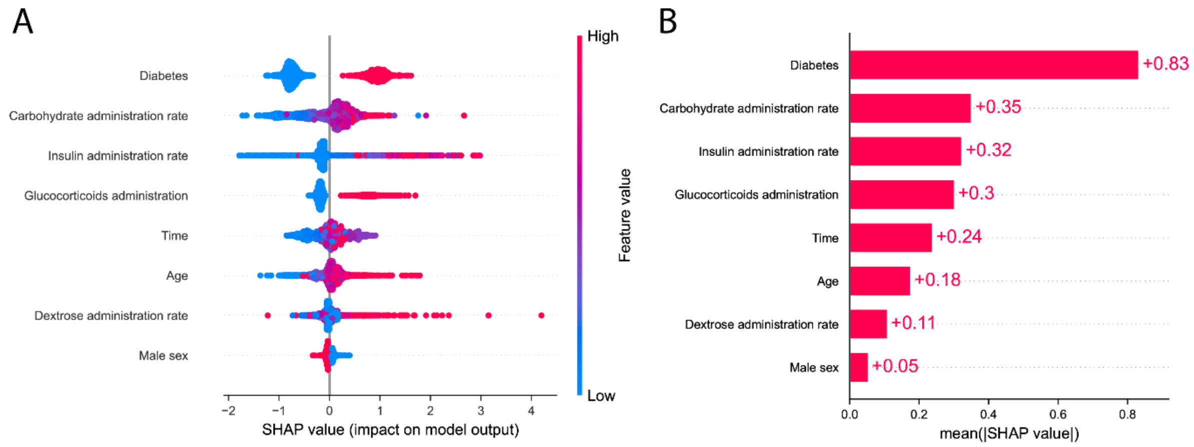


**Supplementary Figure S5: SHAP analysis of XGBoost regression model.** (A) SHAP Summary plot. Each dot represents an individual glucose measurement and is colored according to the value of the feature. Red represents a higher feature value, blue represent a lower value. In categorical features, the red color indicates the presence of the corresponding variable, while blue represents the absence. The horizontal location is of dot depicts whether it corresponds with a higher or lower prediction of the glucose levels (SHAP value). A sample of 5000 randomly selected features was used to in this SHAP analysis. (B) SHAP Global bar plots. For each feature, the global importance is indicated by taking the mean of absolute SHAP values (from panel A) for that feature over all selected samples.

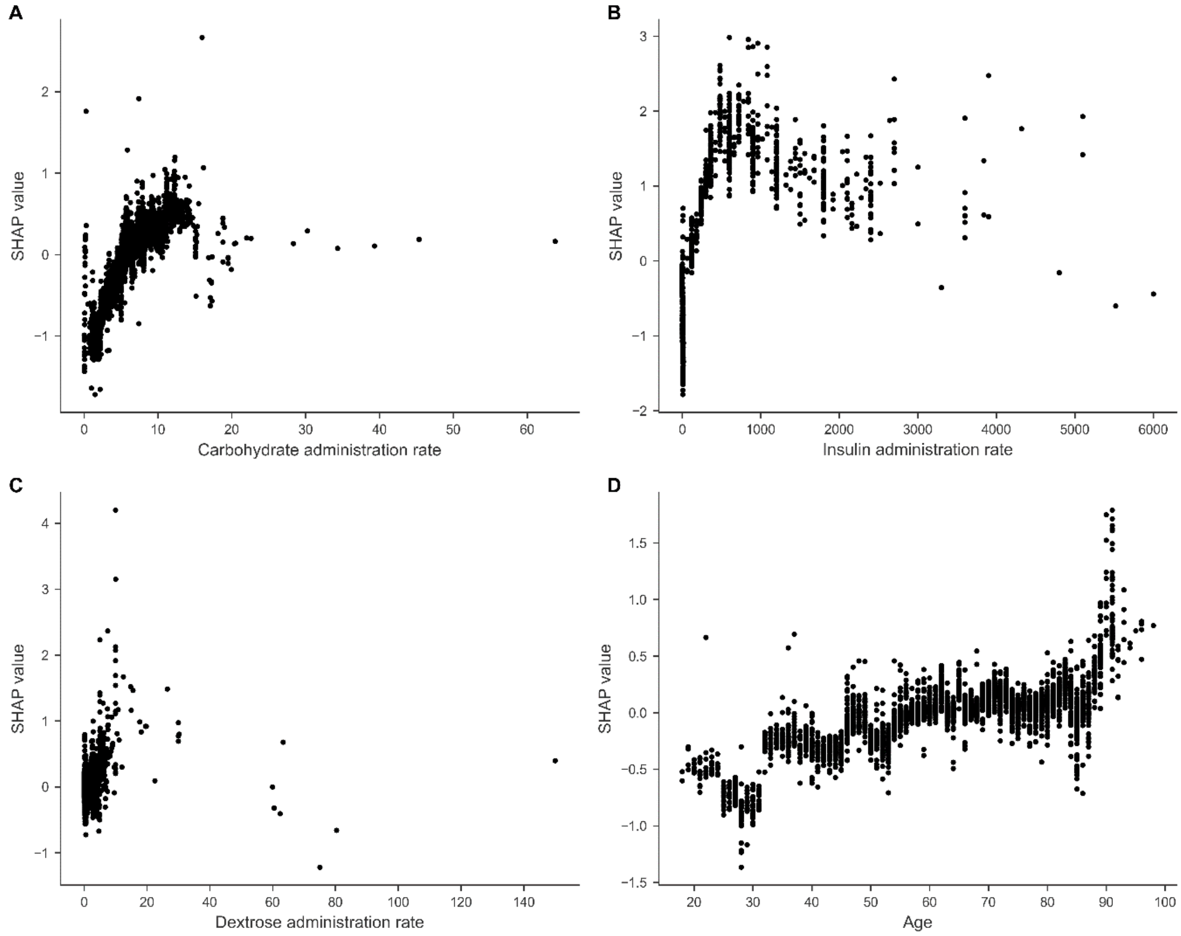


**Supplementary Figure S6: SHAP dependence plots of A) Carbohydrate administration rate, B) Insulin administration rate, C) Dextrose administration rate and D) Age.** Each dot represents an individual glucose measurement, with its feature value at the x-axis and its SHAP value at the y-axis. A sample of 5000 randomly selected features was used to in this SHAP analysis. (B) SHAP Global bar plots.
